# COVID-19 era, Preventive effect of no going out against co-infection of the seasonal influenza virus and SARS-CoV-2

**DOI:** 10.1101/2020.09.27.20202739

**Authors:** Takuma Hayashi, Nobuo Yaegashi, Ikuo Konishi

## Abstract

In the situation where expansion of coronavirus infectious disease-2019 (COVID-19) does not stop, there is concern about co-infection of people with the seasonal influenza infections from late autumn to winter 2020. Therefore, the importance of supplying vaccines against the seasonal influenza has been pointed out all over the world. As an example in Japan, the number of people infected with the seasonal influenza, hand-foot-and-mouth disease (HFMD), epidemic keratoconjunctivitis, and pharyngoconjunctival fever (PCF), which are the seasonal infectious diseases in the 2020 season, has decreased remarkably compared to the number of people infected each year. It is believed that the significant reduction in the number of people infected with these seasonal infectious diseases is a result of the pervasive hand washing, wearing masks and maintaining social distance in COVID-19 rea. To examine the correlation between the three factors of the number of people with each seasonal infectious disease, the mask wearing rate, and the outing rate, we created a three-dimensional scatter plot based on these three factors using principal component analysis. Our research findings demonstrated preventive effect of no going out against co-infection with the seasonal influenza and severe acute respiratory syndrome coronavirus 2 (SARS-CoV-2).

## Introduction

Now that the expansion of coronavirus infectious disease-2019 (COVID-19) is not over, people around the world are afraid of coinfection with the seasonal influenza virus and severe acute respiratory syndrome coronavirus 2 (SARS-CoV-2), which is expected to be found winter in 2020. Since the spread of COVID-19, in Japan, measures to prevent the transmission of the virus such as the use of masks and hand washing, ensuring social distance, refraining from going out and canceling large events are widely promoted.^1,2^ In the case of Tokyo, the number of seasonal infectious diseases such as influenza during the current term (2019–2020) is reported to be significantly lower than in the average year (2015–2018).^3^ We examined whether wearing a mask or refraining from going out had a great influence on the incidence of seasonal infectious diseases by a statistical method using the principal component analysis (PCA) load based on a three-dimensional scatter plot. We demonstrated preventive effect of no going out against co-infection with the seasonal influenza and severe acute respiratory syndrome coronavirus 2 (SARS-CoV-2) compared to wearing a mask.

## Method

In approximately 5000 sentinel centers including hospitals and clinics, the number of persons with the seasonal infectious diseases (seasonal influenza, hand-foot-and-mouth disease, epidemic keratoconjunctivitis) and those infected with SARS-CoV-2 in Tokyo was calculated by the National Institute of Infectious Diseases and Tokyo Metropolitan Infectious Diseases Surveillance Center.^3,4^ The number of people with various seasonal infectious diseases was informed in weekly reports (Week 1–53 of the same year) (Supplementary data).^3^

The mask wearing rate in Tokyo from March–August 2020 was calculated by YouGov international COVID-19 tracker Nippon Research Center Ltd (Tokyo, Japan). The outing rate in Tokyo from March–August 2020 was calculated by the Ministry of Land, Infrastructure, Transport, and Tourism and 2020 Agoop Corp. (Tokyo, Japan).^3^ To examine the correlation between the three factors of the number of people with each seasonal infectious disease, the mask wearing rate, and the outing rate, we created a three-dimensional scatter plot based on these three factors using Minner3D Enterprise-Miner3D1.m3d (XLSTAT by Addinsoft, Mindware Inc., Okayama, Japan).

We calculated the PCA vector from the three-dimensional scatter plot and calculated the PCA load for the number of affected individuals of each seasonal infectious disease, mask wearing rate, and outing rate.

## Result

In the case of Tokyo, the numbers of people infected with the influenza virus, with hand-foot-and-mouth disease, and epidemic keratoconjunctivitis in the current term (2019–2020) were significantly lower than in the average year (2015–2018) (Figure 1). Therefore, we calculated the amount of PCA load for the number of people affected by each seasonal infectious disease, the mask wearing rate, and the outing rate, and examined the effect on the PCA vector.

**Figure 1.**
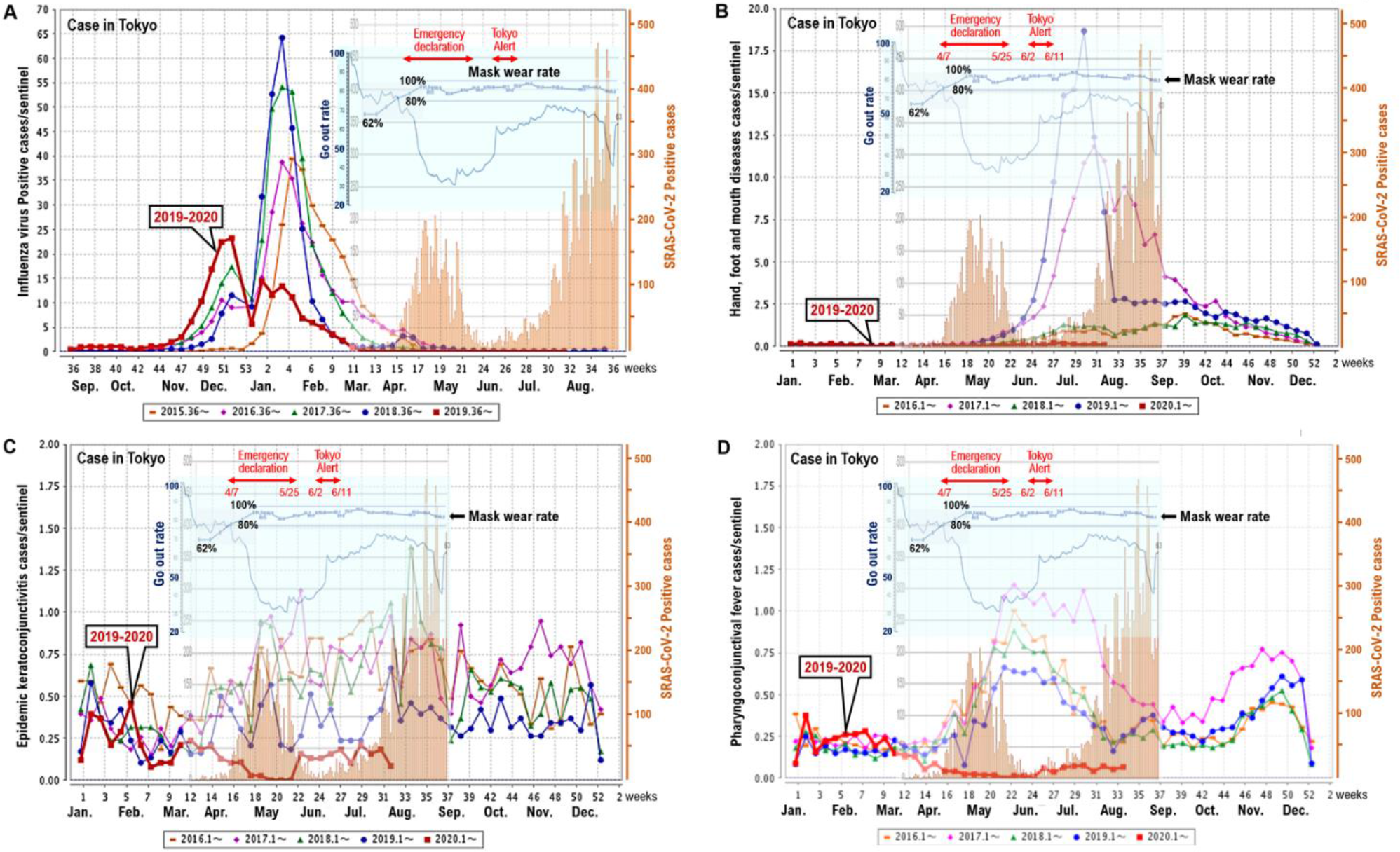
The numbers of patients with the seasonal infectious disease and SARS-CoV-2. The number of peoples with the seasonal infectious diseases, **(A)** influenza virus, **(B)** hand-foot-and-mouth disease, **(C)** epidemic keratoconjunctivitis, **(D)** pharyngoconjunctival fever and the number of peoples infected with SARS-CoV-2 in Tokyo (2015–2020). The mask wearing rate and the go out (outing) rate (March to August, 2020) is indicated in each panel.

three-dimensional scatter plot were created based on these three factors (the numbers of people infected with the seasonal viruses, mask wearing rate, and outing rate) using Minner3D Enterprise-Miner3D1.m3d. From the results of this study, it was clarified that a decrease in mask wearing rate (load −0.566) and an increase in outing rate (load 0.713) influenced the increase in the number of people infected with the seasonal influenza virus (Figure 2A). Particularly, the increase in the outing rate has a strong influence on the increase in the number of people infected with the seasonal influenza virus (Figure 2A). An increase in the outing rate (load 0.591) affected the increase in the number of people with hand-foot-and-mouth disease (HFMD) (Figure 2B). It was clarified that the mask wearing rate (load 0.104) did not affect the increase in the number of people with HFMD (Figure 2B). The results of this study further revealed that a decrease in the mask wearing rate (load −0.443) and an increase in the outing rate (loading 0.713) influenced the increase in the number of people with epidemic keratoconjunctivitis (Figure 2C). The increase in the outing rate has a strong influence on the increase in the number of people with the epidemic keratoconjunctivitis (Figure 2C). The results of this study further revealed that a decrease in the mask wearing rate (load −0.526) and an increase in the outing rate (loading 0.690) influenced the increase in the number of people with pharyngoconjunctival fever (PCF) (Figure 2D). The increase in the outing rate has an influence on the increase in the number of people with the PCF (Figure 2D).

**Figure 2.**
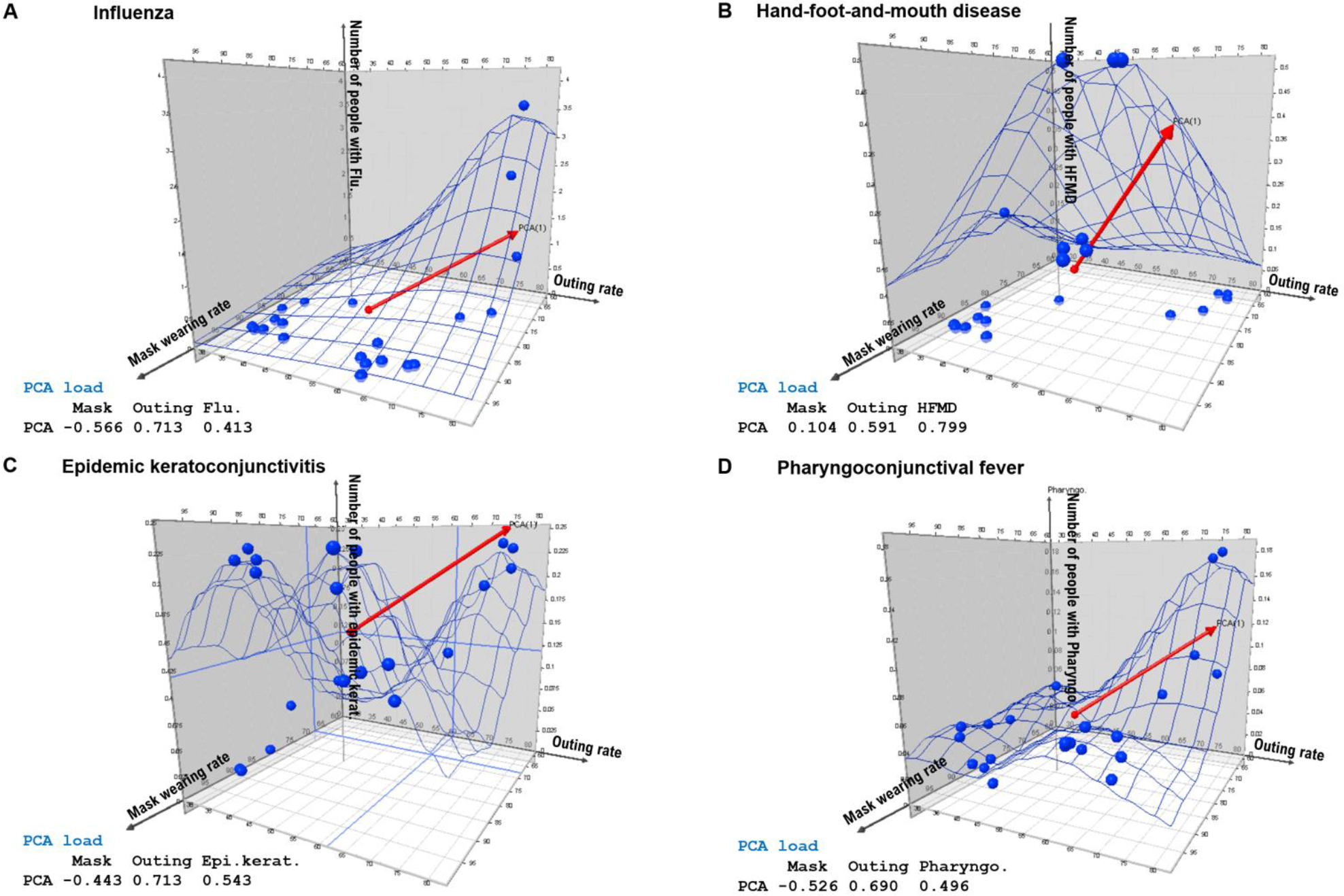
The three-dimensional scatter plot based on these three factors. The three-dimensional scatter plot were created based on these three factors (number of peoples with **(A)** influenza virus, **(B)** hand-foot-and-mouth disease, **(C)** epidemic keratoconjunctivitis, or **(D)** pharyngoconjunctival fever; the mask wearing rate and the go out (outing) rate using Minner3D Enterprise-Miner3D1.m3d. The principal component analysis (PCA) vectors are indicated Red color arrows in the 3D scatter plot data. The PCA loads for the number of affected individuals for each seasonal infectious disease, mask wearing rate, and outing rate are shown in the bottom left corner of each panel.

The number obtained by subtracting the number of seasonal influenza-infected persons in 2019-2020 year from the number of seasonal influenza-infected persons in 2018-2019 year is the effect of suppressing the seasonal influenza infection by social activities such as wearing mice or refraining from going out (Figure 3A). The results of this study further revealed that a decrease in the mask wearing rate (load 0.501) and an increase in the outing rate (loading 0.850) influenced the increase in the number of people with the seasonal influenza virus (Figure 3B). The increase in going out rate has a strong influence on the increase in the number of people with the seasonal influenza virus (Figure 3B).

**Figure 3.**
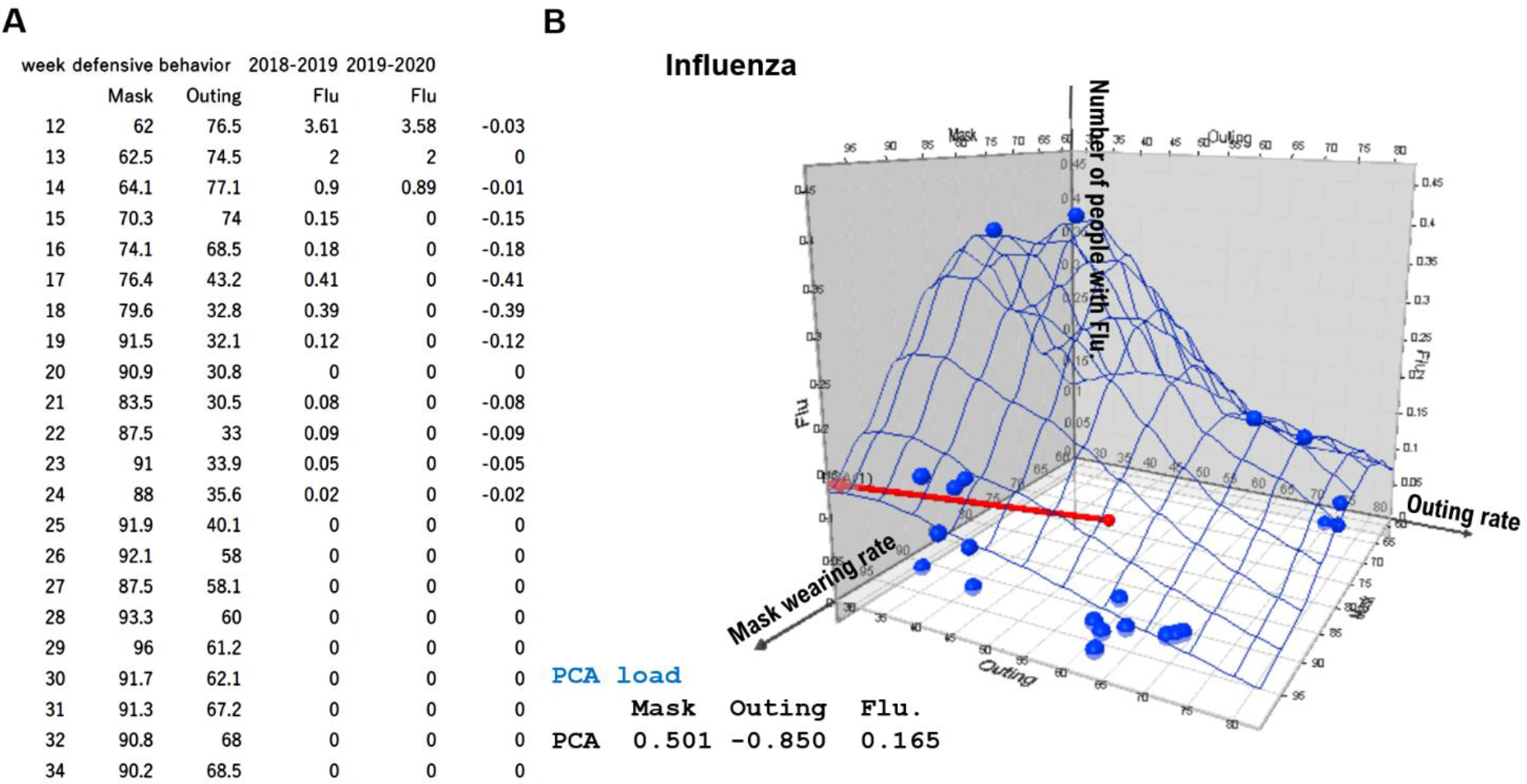
In COVID-19 era, Preventive effect of no going out against co-infection of the seasonal influenza virus. **(A)** The number obtained by subtracting the number of seasonal influenza-infected persons in 2019-2020 year from the number of seasonal influenza-infected persons in 2018-2019 year is the effect of suppressing the seasonal influenza infection by social activities such as wearing mice or refraining from going out. **(B)** The three-dimensional scatter plot were created based on these three factors (number of peoples with influenza virus, the mask wearing rate and the go out (outing) rate using Minner3D Enterprise-Miner3D1.m3d. The principal component analysis (PCA) vectors are indicated Red color arrows in the 3D scatter plot data. The PCA loads for the number of affected individuals for each seasonal infectious disease, mask wearing rate, and outing rate are shown in the bottom left corner of each panel.

In the COVID-19 era, during the period of the state of emergency (April 7, 2020 to May 25, 2020), the number of people going out decreased to about 25.64% compared to the period before the spread of COVID-19 infection (February 2020) in Tokyo (Figure 4). Currently, even in late September 2020, the number of people going out in Tokyo has decreased to about 58.9% compared to the period before the spread of COVID-19 infection (February 2020) in Tokyo (Figure 4). Even during the seasonal influenza epidemic (November 2020 to March 2021), if the number of people going out is reduced to about 60%, the number of people infected with seasonal influenza virus may decrease to about 36% compared to the average year.

**Figure 4.**
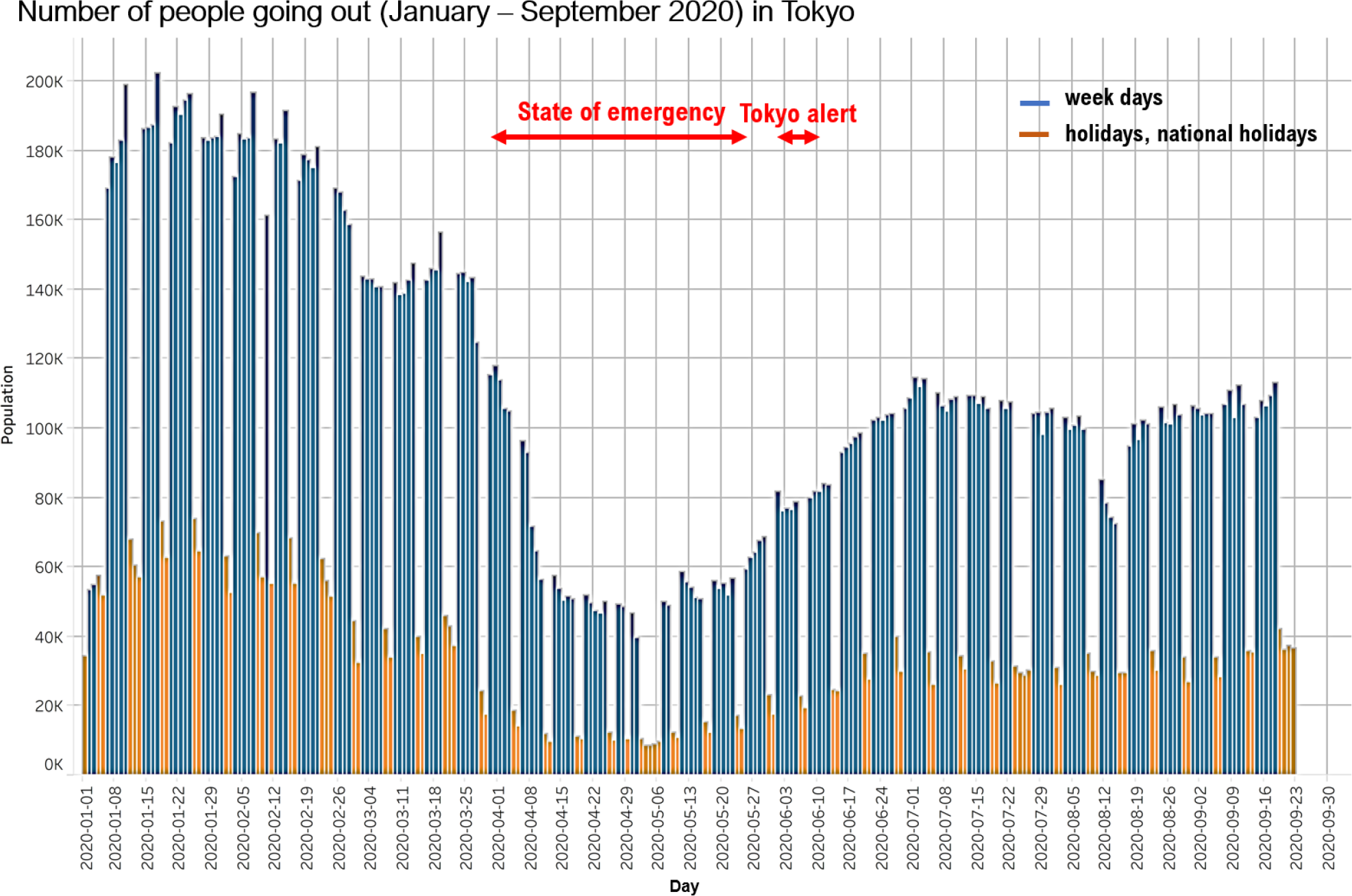
Number of people going out (January – September 2020) in Tokyo. In the COVID-19 era, during the period of the state of emergency (April 7, 2020 to May 25, 2020), the number of people going out decreased to about 25.64% compared to the period before the spread of COVID-19 infection (February 2020) in Tokyo. Currently, even in late September 2020, the number of people going out in Tokyo has decreased to about 58.9% compared to the period before the spread of COVID-19 infection (February 2020) in Tokyo. The graph is adapted from Rf.6.

## Discussion

In the COVID-19 era, people around the world are afraid of coinfection with the seasonal influenza virus and SARS-CoV-2, which is expected to be found winter in 2020. From the results of this analysis, it was predicted that refraining from going out might work strongly as a preventive effect against the seasonal influenza infection. Vaccination, such as the influenza vaccine, is the most important medical strategy to prevent infected individuals from becoming seriously ill due to viral infections.^5^ The results of this study revealed that protective behaviors and social consensus against viral infections are likely to influence the decrease in the number of patients with seasonal infectious disease.

## Data Availability

We show a statement regarding the availability of all data referred to in the manuscript.
https://www.metro.tokyo.lg.jp/english/topics/2020/0128_00.html
http://idsc.tokyo-eiken.go.jp/assets/weekly/2020/33e.pdf
https://stopcovid19.metro.tokyo.lg.jp/en
https://corporate-web.agoop.net/pdf/covid-19/agoop_analysis_coronavirus.pdf

## Data Sharing

Data are available on various websites and have been made publicly available (more information can be found in the first paragraph of the Results section).

## Acknowledgments

We thank Professor Richard A. Young (Whitehead Institute for Biomedical Research, Massachusetts Institute of Technology, Cambridge, MA) for his research assistance. This study was supported in part by grants from the Japan Ministry of Education, Culture, Science and Technology (No. 24592510, No. 15K1079, and No. 19K09840), the Foundation of Osaka Cancer Research, The Ichiro Kanehara Foundation for the Promotion of Medical Science and Medical Care, the Foundation for Promotion of Cancer Research, the Kanzawa Medical Research Foundation, The Shinshu Medical Foundation, and the Takeda Foundation for Medical Science.

## Author Contributions

T.H. performed most of the experiments and coordinated the project; T.H. and N.Y. conceived the study and wrote the manuscript. N.Y. and I.K. gave information on clinical medicine and oversaw the entire study.

## Transparency document

The transparency document associated with this article can be found in the online version at http://

