## supplemental materials for "COVID-19 era, Preventive effect of no going out against co-infection of the seasonal influenza virus and SARS-CoV-2"

| week | defensive behavior | | seasonal infectious disease (2019-2020) | | | |
|---|---|---|---|---|---|---|
| | Mask | Outing | Flu | HFMD | Epidemic | Pharyngo. |
| 12 | 62 | 76.5 | 3.58 | 0 | 0.23 | 0.18 |
| 13 | 62.5 | 74.5 | 2 | 0 | 0.235 | 0.175 |
| 14 | 64.1 | 77.1 | 0.89 | 0 | 0.21 | 0.08 |
| 15 | 70.3 | 74 | 0 | 0 | 0.195 | 0.1 |
| 16 | 74.1 | 68.5 | 0 | 0 | 0.13 | 0.07 |
| 17 | 76.4 | 43.2 | 0 | 0 | 0.09 | 0.07 |
| 18 | 79.6 | 32.8 | 0 | 0.2 | 0.06 | 0.04 |
| 19 | 91.5 | 32.1 | 0 | 0 | 0.013 | 0.05 |
| 20 | 90.9 | 30.8 | 0 | 0 | 0.012 | 0.04 |
| 21 | 83.5 | 30.5 | 0 | 0 | 0.015 | 0.04 |
| 22 | 87.5 | 33 | 0 | 0 | 0.23 | 0.01 |
| 23 | 91 | 33.9 | 0 | 0 | 0.22 | 0.02 |
| 24 | 88 | 35.6 | 0 | 0 | 0.22 | 0.02 |
| 25 | 91.9 | 40.1 | 0 | 0 | 0.21 | 0.01 |
| 26 | 92.1 | 58 | 0 | 0.2 | 0.2 | 0.05 |
| 27 | 87.5 | 58.1 | 0 | 0.2 | 0.229 | 0.04 |
| 28 | 93.3 | 60 | 0 | 0.5 | 0.126 | 0.055 |
| 29 | 96 | 61.2 | 0 | 0.2 | 0.232 | 0.06 |
| 30 | 91.7 | 62.1 | 0 | 0.2 | 0.132 | 0.065 |
| 31 | 91.3 | 67.2 | 0 | 0.5 | 0.141 | 0.03 |
| 32 | 90.8 | 68 | 0 | 0.5 | 0.11 | 0.06 |
| 34 | 90.2 | 68.5 | 0 | 0.2 | 0.12 | 0.045 |
